# The grit personality trait, eating behavior, and obesity among Japanese adults

**DOI:** 10.1101/2024.04.13.24305766

**Authors:** Noriaki Kurita, Takako Maeshibu, Tetsuro Aita, Takafumi Wakita, Hiroe Kikuchi

**Author notes:** Corresponding author: Noriaki Kurita, MD, PhD, FACP Department of Clinical Epidemiology, Graduate School of Medicine Fukushima Medical University 1 Hikarigaoka, Fukushima City, Fukushima 960-1295, Japan. **Clinical Trial Registry number and website where it was obtained** Not applicable.

## Abstract

**Background:** Despite the stigma attributing obesity to a lack of willpower, research on the interrelationships among an individual’s willpower, eating behaviors, and obesity is lacking.

**Objective:** This study aimed to quantify the extent to which multidimensional eating behaviors mediated the association between obesity and grit, which share commonalities with self-control.

**Methods:** This cross-sectional study involved Japanese adults across a wide range of age groups. Grit was measured using an 8-item short grit scale. Multidimensional eating behaviors were measured using the Japanese version of the 21-item Three-Factor Eating Questionnaire-R21, comprising uncontrolled eating, emotional eating, and cognitive restraint. Obesity was defined as a body mass index ≥25.0 kg/m2, which is the World Health Organization’s cutoff specific to Asian populations in the Asia-Pacific region. A series of logistic regression models were created to analyze the association between grit and obesity with and without eating behaviors. Mediation analyses using the Karlson Holm Breen method were performed to determine whether eating behavior mediated this association.

**Results:** Of the 1641 adults, 26.8% were obese. A higher grit level was associated with a lower likelihood of obesity, less uncontrolled and emotional eating, and higher cognitive restraint. Uncontrolled and emotional eating fully mediated the association between grit and obesity, whereas cognitive restraint only partially mediated this association.

**Conclusions:** The inverse association between grit and obesity was mediated by multidimensional eating behaviors. Identifying impairments in eating behaviors, rather than focusing on an individual’s lack of willpower, may contribute to dispelling the aforementioned stigma and facilitating a dialogue for the prevention and management of obesity.

## Introduction

Obesity is a metabolic disease that increases the risk of cardiovascular disease (1) and its prevalence continues to worsen worldwide. From 1975 to 2014, the worldwide mean body mass index (BMI) increased by 2.5 kg/m^2^ and 2.3 kg/m^2^ for men and women, respectively (2). Japan showed a similar trend, particularly among men (3,4). The continuous efforts of individuals to change dietary behavior and effective motivational support from primary care providers are major factors that help individuals prevent obesity-related complications and improve their well-being (5,6). Therefore, primary care providers should assess physiological sensations related to eating, restraint behaviors, and emotion-related behaviors (6). However, rather than providing support, primary care providers may attribute the individual’s unsuccessful continuous management of obesity to their poor willpower or lack of self-control (7), and, thereby, fail to establish a constructive and helpful relationship (6). Despite these persistent stigmas, there is insufficient evidence to understand the interrelationships among individuals’ willpower, eating behavior, and obesity.

Grit personality trait, defined as the ability to maintain commitment to long-term goals despite challenges (8), is associated with healthy eating behaviors and a low prevalence of obesity. However, several cross-sectional studies demonstrating an association between high grit and low BMI or overweight among the US adult population failed to account for differences in exercise habits and eating behaviors, as acknowledged by the authors (9,10). In addition, a study demonstrating an association between high grit, regular eating, and healthy food choices did not address the involvement of eating with negative emotions or hunger (11). Multi-dimensional eating behaviors, including emotional eating (EE: i.e., overeating associated with negative emotions), can be separately assessed using the Three-Factor Eating Questionnaire (TFEQ), along with uncontrolled eating (UE, i.e., general difficulties in regulating eating, including hunger) and cognitive restraint (CR, i.e., conscious restriction of food intake to control weight) (12,13). However, only one study that examined the association between grit and eating behavior, as assessed by the TFEQ, measured only cognitive restraint and not the association in with emotional and uncontrolled eating (14). More importantly, to date, there is a lack of research on the extent to which multidimensional eating behaviors mediate the relationship between low grit and obesity.

Therefore, this study aimed to examine the interrelationships among grit, multidimensional eating behaviors, and obesity in a wide-ranged population of adult Japanese men and women. Furthermore, we quantified the extent to which multidimensional eating behaviors mediate the association between grit and obesity. If the association is mostly mediated by eating behaviors, it would provide a basis for those engaged in obesity management as well as primary care providers to proactively devise ways to assess individuals’ eating behaviors and formulate specific individualized strategies, instead of unjustly blaming obese individuals for having poor willpower.

## Methods

### Setting and Participant Selection

This cross-sectional online survey was approved by the Institutional Review Board at Fukushima Medical University (ippan2022-210). It was a panel survey involving Japanese adults aged ≥20 years, assisted by a web-based company (Cross Marketing, Shinjuku-ku, Tokyo, Japan). The target sample size was 1,500 because of the project’s budget constraints. The respondents were recruited based on stratified groups to sample men and women, older adults (≥65 years) and young adults, and individuals with and without obesity. (Supplementary Item 1) Participants were offered incentive points that could be redeemed for cash, gift certificates, or mileage. Participants answered an online questionnaire prepared by the company. The response data were collected between January 26 and 31, 2023, and stored on the company’s server. The participants were instructed to respond only once. Only those who provided informed consent completed the questionnaire.

### Designing Screener Items

To assess the presence of careless participants (15), five “screener” items were created to identify and exclude them from our analyses. Specifically, we excluded respondents with inappropriate entries of 1) age or 2) gender or extreme values for 3) height, 4) weight, or 5) completion time <5 minutes (16,17). (Supplementary Item 2)

### Grit

The exposure in this study was grit personality trait assessed using the Japanese version of the eight-item Short Grit Scale (8,18). The concept of grit comprises two aspects: a passion for and perseverance toward long-term goals (19). The former represents consistency of interest (e.g., maintaining focus on a project that will take more than a few months to complete), and the latter represents perseverance in the face of specific setbacks (e.g., not being discouraged by setbacks, conquering them, and finishing a project). Respondents were instructed to rate each item on a Likert scale ranging from 1 (“Not at all like me”) to 5 (“Very much like me”). The scores for the four negatively worded items were then reversed, and the average of all items was used as the score. The alpha coefficient for the Short Grit Scale was 0.74. The construct validity of the scale was verified using confirmatory factor analysis (18).

### Obesity

The outcome of this study was obesity defined as a BMI ≥ 25.0 kg/m2. This cutoff is regarded as specific to Asians living in the Asia-Pacific region by the World Health Organization (20) and is also supported by the Japan Society for the Study of Obesity (21). This cut-off has been established as the threshold for an increased risk of diabetes and hypertension in Japanese and other Asian populations (20,21). BMI was calculated as weight (kg) divided by height (in meters) squared using self-reported weight and height data.

### Eating behavior: the Three-Factor Eating Questionnaire-R21

The mediator in this study was multifaceted eating behavior that was measured using the Japanese version of the 21-item Three-Factor Eating Questionnaire-R21 (TFEQ-R21) scale developed by Cappelleri and Karlsson (22). After obtaining permission from the developer (Karlsson), two physicians with experience in scale development (N.K. and H.K.) translated the scale into Japanese. Next, it was back-translated into English by two bilingual translators (one American and one Canadian), and necessary revisions were made to the translation by comparing the wordings with the original. Finally, the back-translated and translated versions were sent to the original author, and the final version was approved (see **Supplementary Table 1**).

Before answering the TFEQ-R21, participants were instructed as follows: “This section contains statements and questions about eating behavior and the feeling of hunger. Read each statement carefully and select the option that best applies to you.” Among the 21 translated items of the TFEQ-R21, participants were instructed to respond to items 1–20 on a Likert scale ranging from 1 to 4 and to item 21 on an eight-point numerical rating scale. Responses to each item were assigned a score between one and four. Before calculating the domain scores, items 1–16 were reverse coded, and item 21 was recoded as follows: scores 1–2 as 1, 3–4 as 2, 5–6 as 3, and 7–8 as 4 (22). Domain scores were then calculated as the mean of all items in each domain or as a transformed score ranging from 0 to 100, where the sum of all items was subtracted from the lowest possible raw score, divided by the range of possible raw scores, and multiplied by 100. The latter method was used in the present study. Higher scores for CR (6 items), UE (9 items), and EE (6 items) indicated greater CR, UE, and EE.

### Other Survey Variables

A detailed description of item selection is provided in the Supplementary Material (**Supplemental Item 3**). The Japanese version of the Dutch Eating Behavior Questionnaire (DEBQ) is a 33-item instrument comprising three factors: “emotional eating,” “external eating,” and “restrained eating” (23,24). Respondents were instructed to rate each item on a Likert scale ranging from 1 (never) to 5 (very often). The average of the items was used as the score for each of the three domains (23). Higher scores indicated that the construct applied better. The alpha coefficients for each domain were as follows: emotional eating, 0.95; external eating, 0.73; restrained eating, 0.87 (24).

Demographic characteristics such as age, sex, education level, total household income, and marital status; health behaviors such as exercise habits, smoking history, and alcohol consumption; and, non-communicable diseases (diabetes, cancer, kidney disease, stroke, congestive heart failure, chronic lung disease, eating disorders, depression, and other psychiatric disorders) were used as covariates.

### Statistical Analysis

Psychometric analyses were performed using R version 4.1.2, the psych package version 2.2.3, and the lavaan package version 0.6-11. All other analyses were performed using Stata/SE, version 17 (Stata Corp., College Station, TX, USA). Respondents’ characteristics are summarized as means and standard deviations and 5th and 95th percentiles for continuous variables and as frequencies and proportions for categorical variables.

For the TFEQ-R21, three-factor confirmatory factor analyses were conducted to examine the goodness-of-fit of the three-factor model. As an exception for this particular analysis, recoded raw scores were used (items 1–16 and item 21). Acceptable criteria for the model fit are a comparative fit index (CFI) ≥ 0.90 and a root mean square approximation error (RMSEA) ≤ 0.08 (25). Acceptable standardized loadings were set at ≥ 0.3 (26). The distribution of responses was evaluated at the item level to identify any significant floor or ceiling effects (>50% of the responses at the lower or upper end of the score) (22). The internal consistency reliability of each domain was assessed by Cronbach’s α and McDonald’s ω coefficients (27). Furthermore, construct validity was examined by testing the correlations between TFEQ-R21 and DEBQ domain scores and BMI. Spearman correlation coefficient was calculated to test these correlations. A detailed description of their hypothesized correlations with the TFEQ-R21 is provided in the Supplementary Material (**Supplemental Item 4**).

Mediation analyses was conducted to examine whether multidimensional eating behavior variables mediated the association between grit and obesity. **Figure 1** illustrates the conceptual framework of this analysis. First, a series of regression models were fitted to informally assess the mediators by observing relative changes in the magnitude of the parameters capturing the association between the exposure variable (i.e., grit) and obesity. Specifically, the association between grit and eating behavior was analyzed using general linear models adjusted for the covariates (see **Figure 1**, path A), and the association between grit and obesity with and without eating behavior was analyzed using logistic regression models adjusted for the covariates (without eating behavior: **Figure 1**, path C; with eating behavior: paths C’ and B). Next, a series of mediation analyses was performed to determine whether eating behavior mediates the relationship between grit and obesity using Stata’s Karlson Holm Breen (KHB) command (28).

**Figure 1:**
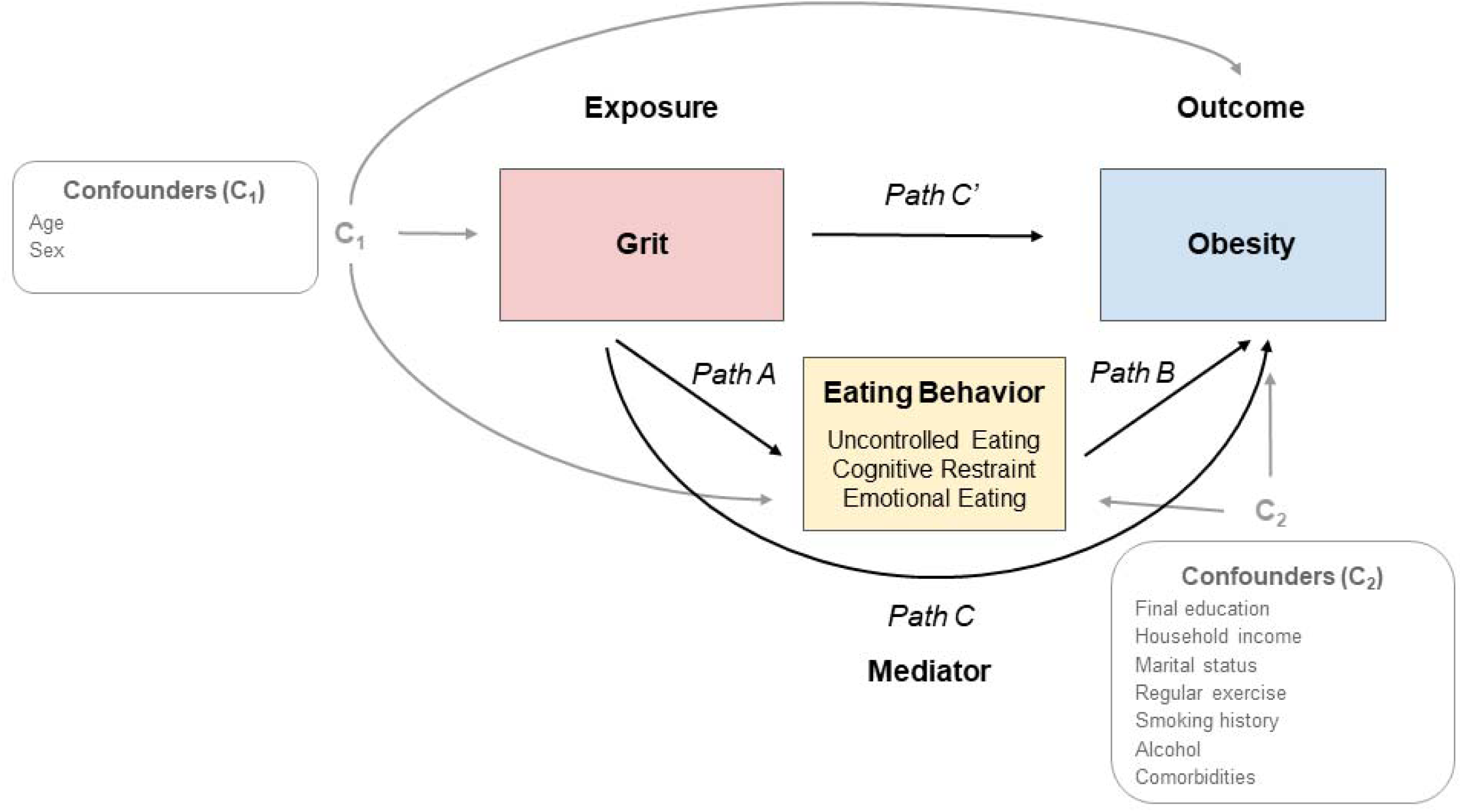
Analytic framework. Path C represents the total effect of grit on obesity. Paths A and B represent the indirect effect via eating behavior. Path C’ represents the indirect effect of grit on obesity controlled for eating behavior.

The KHB method estimates the total, direct, and indirect effects in nonlinear probability models such as logistic regression models (28). Mediation analyses allowed us to estimate the effect of the independent variable (i.e., grit, direct effect) after excluding the effect of the mediators (i.e., eating behavior), the effect explained by the mediators (i.e., indirect effect), and the proportion of the effect mediated by the mediators. The association between grit and obesity was considered completely mediated if the association was no longer evident after controlling for eating behavior and partially mediated if the association persisted (29). The aforementioned series of analyses were performed separately for the UE, CR, and EE domains of the TFEQ-R21. In the sensitivity analyses, the total effect of grit on obesity was decomposed into direct and multiple indirect effects using the KHB method, assuming that exercise habits, smoking, and alcohol consumption, in addition to eating behaviors, were also mediators (28). No data were missing because all items in our online survey had to compulsorily be filled. Statistical significance was set at P < 0.05 for all analyses.

## Results

Overall, there were 2155 participants. After excluding 514 individuals due to carelessness or entering extreme values, 1641 individuals were included in the primary analysis (**Figure 2**).

**Figure 2:**
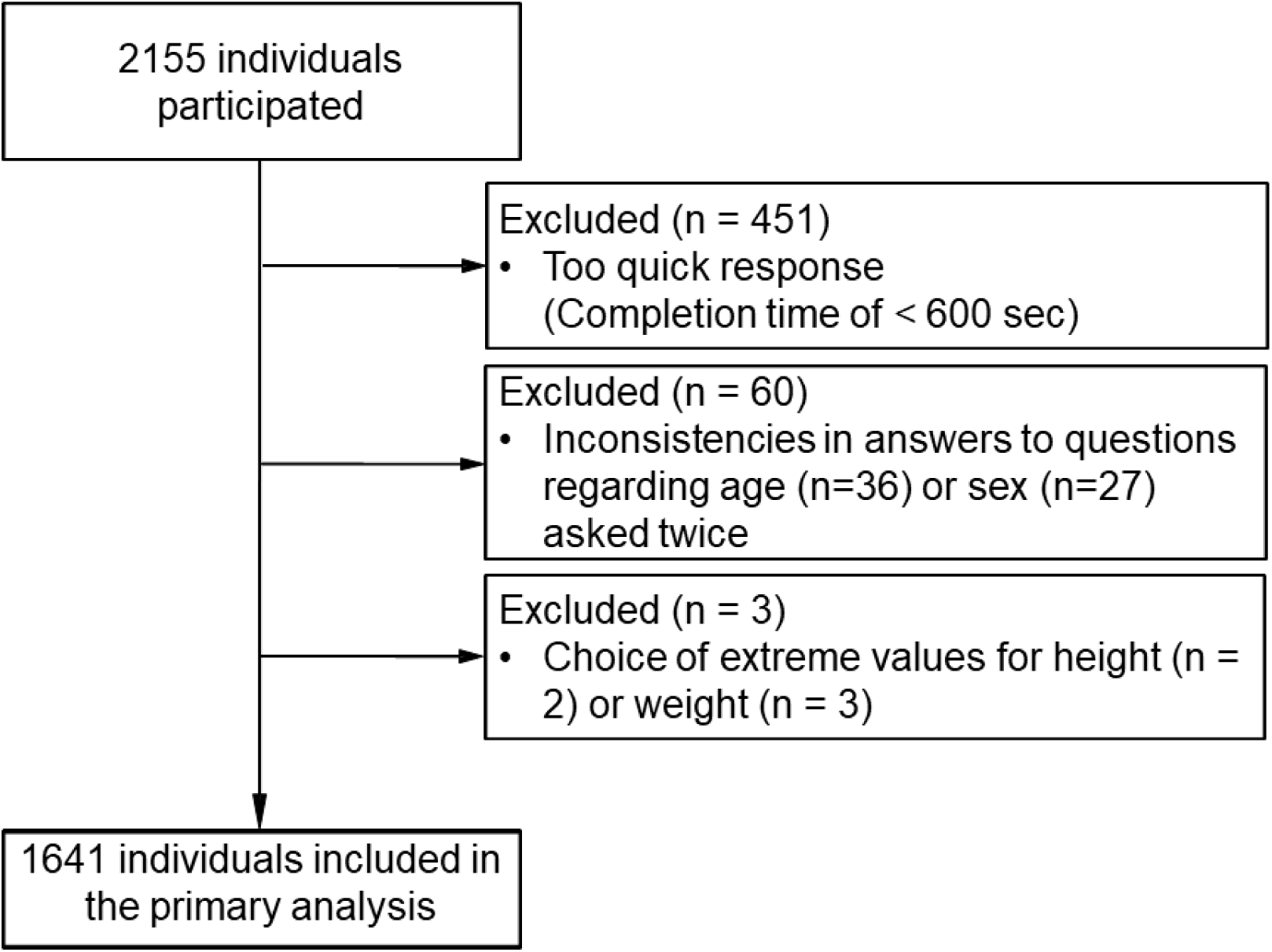
**Flow of the study**

### Participant characteristics

The mean age was 60.6 years, and 786 (47.9%) participants were men (**Table 1**). Diabetes was the most common comorbidity (18%), followed by depression (12.5%) and malignant disease (11.5%). Thirty-one percent participants reported exercising regularly, 19.8% reported consuming alcohol daily, and 18.1% reported smoking. The mean ± SD (5^th^–95th percentile) BMI was 23.0 ± 4.7 (17.2–31.8) kg/m^2^, and 439 (26.8%) participants were obese.

**Table 1.** Participant characteristics by obesity status (N = 1641)

|  | **Obesity** |  | **Total** |
| --- | --- | --- | --- |
|  | *No*<br>n = 1202 | *Yes*<br>n = 439 | N = 1641 |
| <u>Demographics</u> |  |  |  |
| Age, y | 61.1 (13.1) | 59.2 (12) | 60.6 (12.9) |
| Men, n (%) | 549 (45.7 %) | 237 (54 %) | 786 (47.9 %) |
| Education, n (%) |  |  |  |
| Junior high school | 31 (2.6 %) | 18 (4.1 %) | 49 (3 %) |
| High school | 378 (31.5 %) | 161 (36.7 %) | 539 (32.9 %) |
| Professional training college/College of technology/Junior college | 270 (22.5 %) | 82 (18.7 %) | 352 (21.5 %) |
| University | 481 (40 %) | 160 (36.5 %) | 641 (39.1 %) |
| Graduate school | 42 (3.5 %) | 18 (4.1 %) | 60 (3.7 %) |
| Household income, n (%) |  |  |  |
| <1,000,000 yen | 97 (8.1 %) | 41 (9.3 %) | 138 (8.4 %) |
| 1,000,000–<5,000,000 yen | 660 (54.9 %) | 242 (55.1 %) | 902 (55 %) |
| 5,000,000–<10,000,000 yen | 326 (27.1 %) | 123 (28 %) | 449 (27.4 %) |
| >10,000,000 yen | 119 (9.9 %) | 33 (7.5 %) | 152 (9.3 %) |
| Marital status, n (%) |  |  |  |
| Unmarried | 227 (18.9 %) | 110 (25.1 %) | 337 (20.5 %) |
| Married | 791 (65.8 %) | 258 (58.8 %) | 1049 (63.9 %) |
| Divorced | 116 (9.7 %) | 48 (10.9 %) | 164 (10 %) |
| Widowed | 68 (5.7 %) | 23 (5.2 %) | 91 (5.6 %) |
| <u>Comorbidities</u> |  |  |  |
| Diabetes, n (%) | 139 (11.6 %) | 157 (35.8 %) | 296 (18 %) |
| Malignancy, n (%) | 139 (11.6 %) | 50 (11.4 %) | 189 (11.5 %) |
| Renal disease, n (%) | 46 (3.8 %) | 32 (7.3 %) | 78 (4.8 %) |
| Stroke, n (%) | 24 (2 %) | 14 (3.2 %) | 38 (2.3 %) |
| Congestive heart failure, n (%) | 7 (0.6 %) | 5 (1.1 %) | 12 (0.7 %) |
| Chronic lung disease, n (%) | 21 (1.8 %) | 8 (1.8 %) | 29 (1.8 %) |
| Eating disorder, n (%) | 24 (2 %) | 16 (3.6 %) | 40 (2.4 %) |
| Depression, n (%) | 96 (8 %) | 109 (24.8 %) | 205 (12.5 %) |
| Other mental disorders, n (%) | 74 (6.2 %) | 82 (18.7 %) | 156 (9.5 %) |
| <u>Psycho-behavioral characteristics</u> |  |  |  |
| Regular exercise, n (%) | 417 (34.7 %) | 104 (23.7 %) | 521 (31.8 %) |
| Smoking, n (%) | 212 (17.6 %) | 85 (19.4 %) | 297 (18.1 %) |
| Alcohol, n (%) |  |  |  |
| Rarely (cannot) drink | 593 (49.3 %) | 263 (59.9 %) | 856 (52.2 %) |
| Sometimes | 366 (30.5 %) | 94 (21.4 %) | 460 (28 %) |
| Everyday | 243 (20.2 %) | 82 (18.7 %) | 325 (19.8 %) |
| Grit-S, pt | 3.1 [2.9,3.5] | 3 [2.6,3.4] | 3.1 [2.8,3.5] |
| Uncontrolled eating, pt | 38.9 [22.2,50] | 44.4 [33.3,55.6] | 38.9 [22.2,50] |
| Cognitive restraint, pt | 25.9 [11.1,37] | 37 [25.9,51.9] | 29.6 [14.8,40.7] |
| Emotional eating, pt | 11.1 [0,33.3] | 33.3 [11.1,44.4] | 16.7 [0,33.3] |

Descriptive Statistics and Psychometric Testing of the Eating Behavior Scale (TFEQ-R21) The model fit of our data was acceptable (RMSEA = 0.069, CFI = 0.924; **Supplementary Table 2**), and the scales had good reliability (Cronbach’s alpha coefficients: 0.79, CR domain; 0.89, UE domain; 0.92, EE domain). The smallest standardized loading was 0.31 for item 17, which was deemed acceptable considering the aforementioned model fit and reliability (**Supplementary Figure 1**).

The mean ± SD for the CR, UE, and EE domain scores were 38.5 ± 20.0, 30.1 ± 19.3, and 22.4 ± 22.2, respectively. At the item level, three items comprising the EE domain had percentages at the lowest scores being > 50 %; however, no item-level floor effect was observed in the obesity population. (**Supplementary Table 3**).

Correlations in the UE domain were moderate for “external eating” and “emotional eating” and negligible for “restrained eating” in the DEBQ (**Supplemental Table 4**). Correlations in the CR domain were moderate with “restrained eating” but almost negligible with “ external eating” and “emotional eating”. Correlations in the EE domain were strong with “emotional eating,” moderate with “external eating,” and weak with “restrained eating. The UE, CR, and EE domains demonstrated weak positive correlations with BMI (**Supplementary Table 4**). These results support the construct validity of the TFEQ21.

### Association between grit and eating behavior

The median grit score was 3.1 (IQR: 2.8–3.5). **Table 2** shows the associations among eating behavior, grit score, and patient characteristics. The grit score was inversely associated with UE (per 1-point increase: -7.63 [95%CI: -9.13, -6.12]) and EE (per 1-point increase: -6.98 [95%CI: - 8.73, -5.23]). In contrast, the grit score was positively associated with CR (per 1-point increase: 2.51 [95%CI: 0.85, 4.18]). Smoking, diabetes, renal disease, eating disorders, and depression were positively associated with UE, whereas age, male sex, and chronic lung disease were inversely associated. Bereavement, eating disorders, depression, and other psychiatric disorders were positively associated with EE, whereas age, male sex, and chronic lung disease were inversely associated with EE. Diabetes and depression were positively associated with CR, whereas smoking was inversely associated.

**Table 2.**
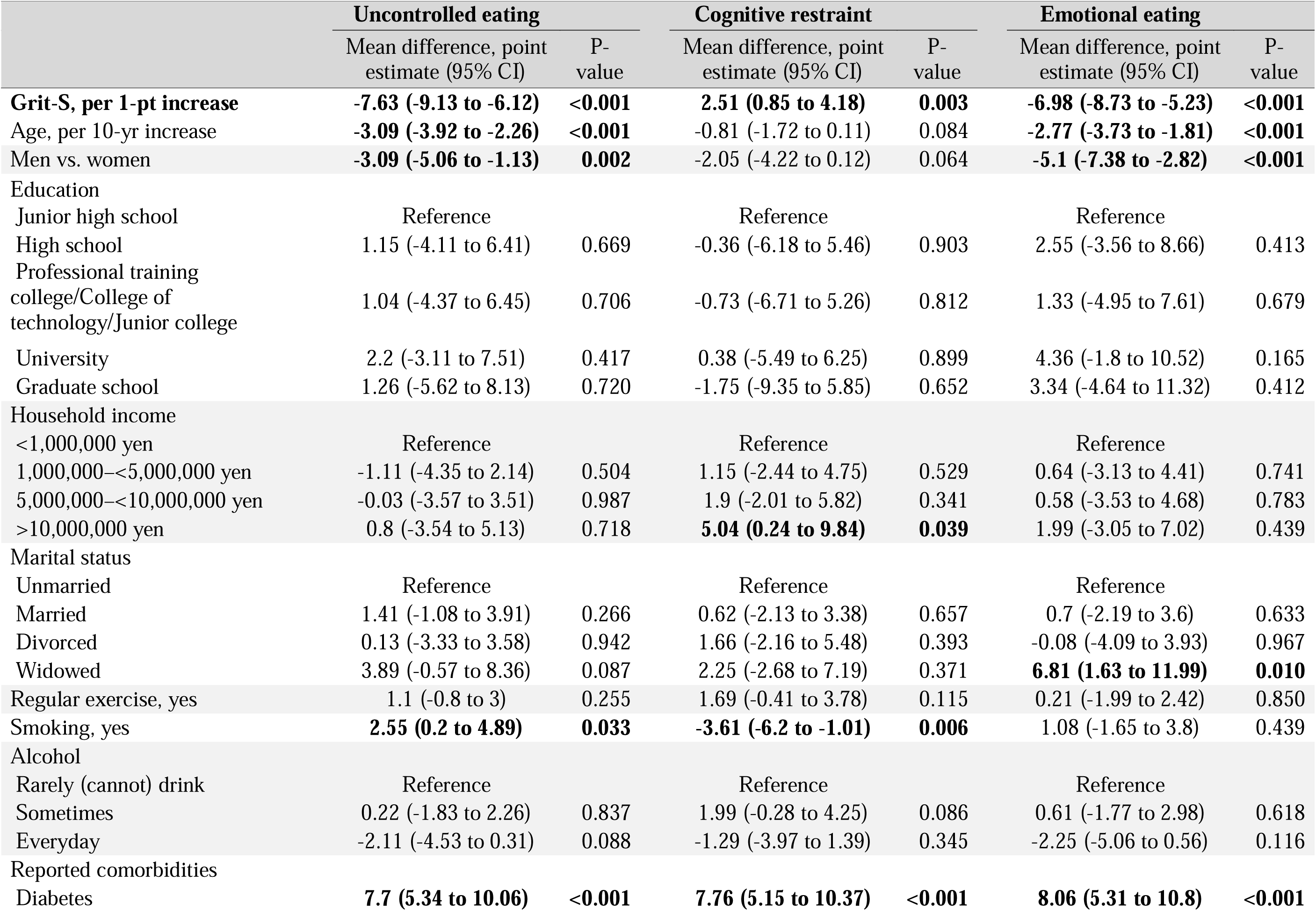
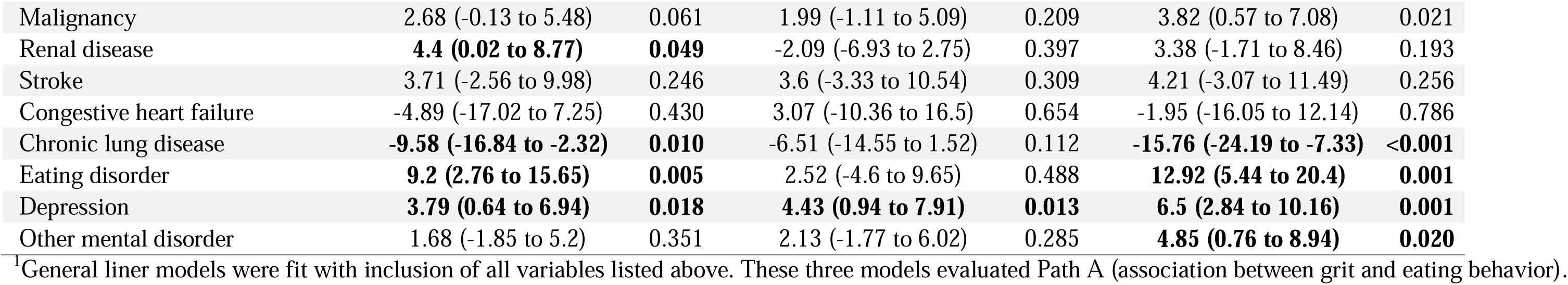
Associations of eating behavior with grit personality and covariates^1^ (N= 1641)

|  | Uncontrolled eating |  | Cognitive restraint |  | Emotional eating |  |
| --- | --- | --- | --- | --- | --- | --- |
|  | Mean difference, point estimate (95% CI) | P-value | Mean difference, point estimate (95% CI) | P-value | Mean difference, point estimate (95% CI) | P-value |
| **Grit-S, per 1-pt increase** | **-7.63 (-9.13 to -6.12)** | **<0.001** | **2.51 (0.85 to 4.18)** | **0.003** | **-6.98 (-8.73 to -5.23)** | **<0.001** |
| Age, per 10-yr increase | **-3.09 (-3.92 to -2.26)** | **<0.001** | -0.81 (-1.72 to 0.11) | 0.084 | **-2.77 (-3.73 to -1.81)** | **<0.001** |
| Men vs. women | **-3.09 (-5.06 to -1.13)** | **0.002** | -2.05 (-4.22 to 0.12) | 0.064 | **-5.1 (-7.38 to -2.82)** | **<0.001** |
| Education |  |  |  |  |  |  |
| Junior high school | Reference |  | Reference |  | Reference |  |
| High school | 1.15 (-4.11 to 6.41) | 0.669 | -0.36 (-6.18 to 5.46) | 0.903 | 2.55 (-3.56 to 8.66) | 0.413 |
| Professional training college/College of technology/Junior college | 1.04 (-4.37 to 6.45) | 0.706 | -0.73 (-6.71 to 5.26) | 0.812 | 1.33 (-4.95 to 7.61) | 0.679 |
| University | 2.2 (-3.11 to 7.51) | 0.417 | 0.38 (-5.49 to 6.25) | 0.899 | 4.36 (-1.8 to 10.52) | 0.165 |
| Graduate school | 1.26 (-5.62 to 8.13) | 0.720 | -1.75 (-9.35 to 5.85) | 0.652 | 3.34 (-4.64 to 11.32) | 0.412 |
| Household income |  |  |  |  |  |  |
| <1,000,000 yen | Reference |  | Reference |  | Reference |  |
| 1,000,000—<5,000,000 yen | -1.11 (-4.35 to 2.14) | 0.504 | 1.15 (-2.44 to 4.75) | 0.529 | 0.64 (-3.13 to 4.41) | 0.741 |
| 5,000,000—<10,000,000 yen | -0.03 (-3.57 to 3.51) | 0.987 | 1.9 (-2.01 to 5.82) | 0.341 | 0.58 (-3.53 to 4.68) | 0.783 |
| >10,000,000 yen | 0.8 (-3.54 to 5.13) | 0.718 | **5.04 (0.24 to 9.84)** | **0.039** | 1.99 (-3.05 to 7.02) | 0.439 |
| Marital status |  |  |  |  |  |  |
| Unmarried | Reference |  | Reference |  | Reference |  |
| Married | 1.41 (-1.08 to 3.91) | 0.266 | 0.62 (-2.13 to 3.38) | 0.657 | 0.7 (-2.19 to 3.6) | 0.633 |
| Divorced | 0.13 (-3.33 to 3.58) | 0.942 | 1.66 (-2.16 to 5.48) | 0.393 | -0.08 (-4.09 to 3.93) | 0.967 |
| Widowed | 3.89 (-0.57 to 8.36) | 0.087 | 2.25 (-2.68 to 7.19) | 0.371 | **6.81 (1.63 to 11.99)** | **0.010** |
| Regular exercise, yes | 1.1 (-0.8 to 3) | 0.255 | 1.69 (-0.41 to 3.78) | 0.115 | 0.21 (-1.99 to 2.42) | 0.850 |
| Smoking, yes | **2.55 (0.2 to 4.89)** | **0.033** | **-3.61 (-6.2 to -1.01)** | **0.006** | 1.08 (-1.65 to 3.8) | 0.439 |
| Alcohol |  |  |  |  |  |  |
| Rarely (cannot) drink | Reference |  | Reference |  | Reference |  |
| Sometimes | 0.22 (-1.83 to 2.26) | 0.837 | 1.99 (-0.28 to 4.25) | 0.086 | 0.61 (-1.77 to 2.98) | 0.618 |
| Everyday | -2.11 (-4.53 to 0.31) | 0.088 | -1.29 (-3.97 to 1.39) | 0.345 | -2.25 (-5.06 to 0.56) | 0.116 |
| Reported comorbidities |  |  |  |  |  |  |
| Diabetes | **7.7 (5.34 to 10.06)** | **<0.001** | **7.76 (5.15 to 10.37)** | **<0.001** | **8.06 (5.31 to 10.8)** | **<0.001** |
| Malignancy | 2.68 (-0.13 to 5.48) | 0.061 | 1.99 (-1.11 to 5.09) | 0.209 | 3.82 (0.57 to 7.08) | 0.021 |
| Renal disease | **4.4 (0.02 to 8.77)** | **0.049** | -2.09 (-6.93 to 2.75) | 0.397 | 3.38 (-1.71 to 8.46) | 0.193 |
| Stroke | 3.71 (-2.56 to 9.98) | 0.246 | 3.6 (-3.33 to 10.54) | 0.309 | 4.21 (-3.07 to 11.49) | 0.256 |
| Congestive heart failure | -4.89 (-17.02 to 7.25) | 0.430 | 3.07 (-10.36 to 16.5) | 0.654 | -1.95 (-16.05 to 12.14) | 0.786 |
| Chronic lung disease | **-9.58 (-16.84 to -2.32)** | **0.010** | -6.51 (-14.55 to 1.52) | 0.112 | **-15.76 (-24.19 to -7.33)** | **<0.001** |
| Eating disorder | **9.2 (2.76 to 15.65)** | **0.005** | 2.52 (-4.6 to 9.65) | 0.488 | **12.92 (5.44 to 20.4)** | **0.001** |
| Depression | **3.79 (0.64 to 6.94)** | **0.018** | **4.43 (0.94 to 7.91)** | **0.013** | **6.5 (2.84 to 10.16)** | **0.001** |
| Other mental disorder | 1.68 (-1.85 to 5.2) | 0.351 | 2.13 (-1.77 to 6.02) | 0.285 | **4.85 (0.76 to 8.94)** | **0.020** |

<sup>1</sup>General liner models were fit with inclusion of all variables listed above. These three models evaluated Path A (association between grit and eating behavior).

### Association of obesity with grit and eating behavior

**Table 3** shows the associations among obesity, grit, and eating behaviors. A higher grit level was associated with a lower likelihood of obesity (grit, per point increase: aOR 0.78 [95%CI: 0.63, 0.97]; model without eating behavior). Furthermore, after adjusting for UE or EE, grit and obesity were not associated. In contrast, when adjusted for CR, the inverse association between grit and obesity was enhanced (grit, per 1-point increase: aOR 0.73 [95%CI: 0.59, 0.91]; model with CR). Higher eating behavior scores were positively associated with obesity (UE, per 10- point increase: aOR 1.39 [95%CI: 1.29, 1.49]; CR, per 10-point increase: aOR 1.17 [95%CI: 1.1, 1.25]; and, EE, per 10-point increase: aOR 1.22 [95 CI: 1.15, 1.29], in respective models including the respective domain).

**Table 3.** Associations of obesity with grit personality, eating behavior, and covariates^1^ (N = 1641)

|  | Model without eating behavior |  | Models with eating behavior |  |  |  |  |  |
| --- | --- | --- | --- | --- | --- | --- | --- | --- |
|  |  |  | Uncontrolled eating |  | Cognitive restraint |  | Emotional eating |  |
|  | Odds ratio, point estimate (95% CI) | P-value | Odds ratio, point estimate (95% CI) | P-value | Odds ratio, point estimate (95% CI) | P-value | Odds ratio, point estimate (95% CI) | P-value |
| Grit-S, per 1-pt increase | **0.78 (0.63 to 0.97)** | **0.023** | 0.99 (0.79 to 1.24) | 0.937 | **0.73 (0.59 to 0.91)** | **0.005** | 0.88 (0.71 to 1.1) | 0.263 |
| **Eating behavior score, per 10-pt increase** |  |  | **1.39 (1.29 to 1.49)** | **<0.001** | **1.17 (1.1 to 1.25)** | **<0.001** | **1.22 (1.15 to 1.29)** | **<0.001** |
| Age, per 10-yr increase | 0.95 (0.85 to 1.07) | 0.401 | 1.06 (0.94 to 1.2) | 0.352 | 0.96 (0 to 1.08) | 0.472 | 1.01 (0 to 1.14) | 0.875 |
| **Men vs. women** | **2.06 (1.56 to 2.73)** | **<0.001** | **2.4 (1.79 to 3.22)** | **<0.001** | **2.18 (1.64 to 2.9)** | **<0.001** | **2.34 (1.75 to 3.13)** | **<0.001** |
| Education |  |  |  |  |  |  |  |  |
| Junior high school | Reference |  | Reference |  |  |  |  |  |
| High school | 1.19 (0.6 to 2.35) | 0.611 | 1.16 (0.58 to 2.34) | 0.675 | 1.24 (0.62 to 2.48) | 0.536 | 1.15 (0.58 to 2.3) | 0.690 |
| Professional training college/College of technology/Junior college | 0.86 (0.42 to 1.74) | 0.671 | 0.82 (0.4 to 1.71) | 0.602 | 0.89 (0.44 to 1.83) | 0.754 | 0.84 (0.41 to 1.72) | 0.627 |
| University | 0.95 (0.48 to 1.89) | 0.889 | 0.87 (0.43 to 1.78) | 0.710 | 0.97 (0.48 to 1.95) | 0.923 | 0.88 (0.43 to 1.77) | 0.712 |
| Graduate school | 0.997 (0.4 to 2.47) | 0.994 | 0.96 (0.38 to 2.45) | 0.940 | 1.06 (0.42 to 2.66) | 0.903 | 0.95 (0.38 to 2.39) | 0.916 |
| Household income |  |  |  |  |  |  |  |  |
| <1,000,000 yen | Reference |  | Reference |  |  |  |  |  |
| 1,000,000—<5,000,000 yen | 1.23 (0.78 to 1.94) | 0.365 | 1.32 (0.83 to 2.11) | 0.247 | 1.21 (0.77 to 1.91) | 0.403 | 1.22 (0.77 to 1.93) | 0.405 |
| 5,000,000—<10,000,000 yen | 1.3 (0.79 to 2.12) | 0.300 | 1.33 (0.8 to 2.21) | 0.274 | 1.28 (0.78 to 2.09) | 0.331 | 1.28 (0.78 to 2.11) | 0.333 |
| >10,000,000 yen | 1.16 (0.62 to 2.15) | 0.639 | 1.14 (0.6 to 2.15) | 0.691 | 1.09 (0.59 to 2.03) | 0.788 | 1.1 (0.59 to 2.06) | 0.769 |
| Marital status |  |  |  |  |  |  |  |  |
| Unmarried | Reference |  | Reference |  |  |  |  |  |
| Married | 0.84 (0.6 to 1.18) | 0.323 | 0.78 (0.55 to 1.11) | 0.164 | 0.83 (0.59 to 1.17) | 0.280 | 0.82 (0.58 to 1.16) | 0.265 |
| Divorced | 0.82 (0.51 to 1.32) | 0.417 | 0.79 (0.49 to 1.29) | 0.353 | 0.78 (0.48 to 1.27) | 0.321 | 0.82 (0.5 to 1.33) | 0.413 |
| Widowed | 0.91 (0.48 to 1.71) | 0.758 | 0.79 (0.41 to 1.51) | 0.476 | 0.88 (0.46 to 1.66) | 0.690 | 0.8 (0.42 to 1.51) | 0.493 |
| **Regular exercise, yes** | **0.69 (0.53 to 0.92)** | **0.010** | **0.64 (0.48 to 0.86)** | **0.002** | **0.67 (0.51 to 0.89)** | **0.005** | **0.68 (0.51 to 0.9)** | **0.007** |
| Smoking, yes | 0.88 (0.64 to 1.22) | 0.451 | 0.8 (0.57 to 1.13) | 0.205 | 0.92 (0.67 to 1.28) | 0.639 | 0.86 (0.62 to 1.19) | 0.367 |
| **Alcohol** |  |  |  |  |  |  |  |  |
| Rarely (cannot) drink | Reference |  | Reference |  |  |  |  |  |
| **Sometimes** | **0.67 (0.5 to 0.9)** | **0.008** | **0.65 (0.48 to 0.88)** | **0.006** | **0.65 (0.48 to 0.87)** | **0.004** | **0.65 (0.48 to 0.89)** | **0.006** |
| Everyday | 0.81 (0.58 to 1.14) | 0.226 | 0.88 (0.62 to 1.24) | 0.456 | 0.82 (0.59 to 1.16) | 0.264 | 0.86 (0.61 to 1.21) | 0.378 |
| Reported comorbidities |  |  |  |  |  |  |  |  |
| **Diabetes** | **5.01 (3.69 to 6.8)** | **<0.001** | **4.29 (3.12 to 5.89)** | **<0.001** | **4.57 (3.35 to 6.22)** | **<0.001** | **4.5 (3.3 to 6.15)** | **<0.001** |
| Malignancy | 0.81 (0.54 to 1.21) | 0.297 | 0.74 (0.49 to 1.12) | 0.151 | 0.78 (0.52 to 1.18) | 0.239 | 0.75 (0.5 to 1.14) | 0.178 |
| Renal disease | 1.62 (0.91 to 2.89) | 0.104 | 1.4 (0.78 to 2.54) | 0.261 | 1.66 (0.93 to 2.97) | 0.088 | 1.53 (0.85 to 2.75) | 0.154 |
| Stroke | 0.92 (0.39 to 2.16) | 0.847 | 0.8 (0.33 to 1.91) | 0.610 | 0.89 (0.38 to 2.1) | 0.797 | 0.79 (0.33 to 1.89) | 0.596 |
| Congestive heart failure | 0.41 (0.08 to 1.98) | 0.266 | 0.44 (0.09 to 2.19) | 0.319 | 0.39 (0.08 to 1.88) | 0.240 | 0.39 (0.08 to 1.95) | 0.254 |
| Chronic lung disease | 0.32 (0.1 to 1.02) | 0.054 | 0.46 (0.14 to 1.48) | 0.195 | 0.34 (0.11 to 1.09) | 0.070 | 0.49 (0.15 to 1.52) | 0.215 |
| Eating disorder | 0.81 (0.34 to 1.94) | 0.634 | 0.61 (0.25 to 1.51) | 0.287 | 0.77 (0.33 to 1.83) | 0.556 | 0.62 (0.25 to 1.52) | 0.296 |
| **Depression** | **2.7 (1.82 to 4.01)** | **<0.001** | **2.59 (1.72 to 3.9)** | **<0.001** | **2.52 (1.69 to 3.75)** | **<0.001** | **2.45 (1.64 to 3.66)** | **<0.001** |
| **Other mental disorder** | **2.06 (1.33 to 3.2)** | **0.001** | **2.01 (1.26 to 3.19)** | **0.003** | **2.02 (1.3 to 3.13)** | **0.002** | **1.93 (1.23 to 3.04)** | **0.004** |

<sup>1</sup>Logistic regression model was fit with inclusion of all variables listed above.
The model without eating behavior evaluated path C' (association between grit and obesity), and the three models with eating behavior informally evaluated paths B (association between eating behavior and obesity) and C (association between grit and obesity after controlling for eating behavior).

### Mediation effect of eating behavior on the relationship between grit and obesity

**Table 4** presents the results of the mediation analyses. Grit was inversely associated with obesity, and UE fully mediated 96.5% of the association. Similarly, EE fully mediated 52.2% of the associations, and CR partially mediated -14.8% of its correlations; in other words, the magnitude of the association of the direct effect was greater in the opposite direction than that of the total effect, indicating that the direct effect was suppressed. Sensitivity analyses examining the overall mediating effects of eating behavior, regular exercise, smoking, and alcohol consumption on the relationship between grit and obesity also revealed a similar degree of mediation (**Supplemental Table 5**).

**Table 4.** Decomposition of the association between grit and obesity into direct and indirect effects using the KHB method^1^ (N = 1641)

|  | Eating behavior as a mediator |  |  |  |  |  |
| --- | --- | --- | --- | --- | --- | --- |
|  | Uncontrolled eating |  | Cognitive restraint |  | Emotional eating |  |
|  | Odds ratio, point estimate (95% CI) | P-value | Odds ratio, point estimate (95% CI) | P-value | Odds ratio, point estimate (95% CI) | P-value |
| Indirect effect | **0.78 (0.72 to 0.84)** | **<0.001** | **1.04 (1.01 to 1.07)** | **0.011** | **0.87 (0.83 to 0.92)** | **<0.001** |
| Direct effect | 0.99 (0.79 to 1.24) | 0.937 | **0.73 (0.59 to 0.91)** | **0.005** | 0.88 (0.71 to 1.1) | 0.263 |
| Total effect | **0.77 (0.62 to 0.96)** | **0.021** | **0.76 (0.62 to 0.95)** | **0.013** | **0.77 (0.62 to 0.96)** | **0.018** |
| % of total effect mediated | 96.5% |  | -14.8% |  | 52.2% |  |

<sup>1</sup>The KHB method was used, which is derived from a linear latent variable model assumed to underlie the logit model and extend the decomposition properties of the linear model to the logit model. This allowed the estimation of the overall, direct, and indirect effects in the logit model (obesity as the dependent variable, grit as the exposure variable, eating behavior as the mediator variable, and the other variables in Table 1 as covariates). This model formally evaluated Path C' (association between grit and obesity), Path B (association between eating behavior and obesity), and Path C (association between grit and obesity after controlling for eating behavior).

## Discussion

Higher grit was associated with a lower likelihood of obesity among Japanese adults. Furthermore, grit was positively associated with cognitive restraint and negatively associated with uncontrolled and emotional eating; these multidimensional eating behaviors mediated the association between grit and obesity. Further research should focus on the abnormalities and causes of mediating eating behaviors rather than ascribing the responsibility of obesity to an individual’s lack of willpower.

Our findings on the interrelationships between grit, eating behaviors, and obesity support previous neuroscientific findings on the grit personality trait and eating behaviors and extend the findings of previous studies on grit and obesity. First, studies in the US demonstrating correlations between grit and low BMI or less likelihood of obesity were limited to young adults and did not account for differences in eating behaviors or exercise habits (9,10). Thus, it is unclear how differences in grit traits generate differences in healthy behaviors and, thus, in body composition. Indeed, our study demonstrated that regular exercise was inversely associated with obesity, independent of grit. Second, the differences in the multidimensional eating behaviors associated with high grit levels corroborate previous research showing the association between grit, brain activity, and eating behavior. Individuals with high grit levels are more likely to engage in a regular diet and healthy food choices (11); they also have greater functional connectivity density in the right dorsolateral prefrontal cortex (DLPFC) (30), which is associated with self-regulation of eating behaviors. Thus, the increased DLPFC activity associated with high grit levels may lead to less uncontrolled and emotional eating and increased cognitive restraint, contributing to the establishment of healthy eating habits. Third, unlike previous studies (9,10), we demonstrated that by isolating the mediating effects of multidimensional eating behaviors, the low likelihood of obesity due to high grit levels can be mostly mediated by the inhibition of uncontrolled or emotional eating.

These findings were further supported by sensitivity analyses, which showed a larger proportion of mediation by eating behavior than that by regular exercise.

Our findings highlight the need for physicians and researchers to focus on differences in grit levels and the presence of mediation through multidimensional eating behaviors to devise strategies to prevent or improve obesity. First, the complete mediating role of unrestricted and emotional eating in the association between high grit levels and low likelihood of obesity indicates the importance of regulating uncontrolled and emotional eating in individuals with low grit levels. Thus, primary care physicians may need to develop measures to assess and improve an individual’s level of self-regulation over time through repeated dialogues about their thoughts, feelings, and behaviors related to eating (6,31). Indeed, mindfulness and cognitive behavioral therapy may contribute to improvements in BMI by inhibiting uncontrolled and emotional eating (31). Second, one should not be bound by incorrect stereotypes that attribute obesity to traits such as lack of willpower (7,32), but rather re-emphasize that, as Stunkard et al. pioneered long ago, obesity is a multifaceted and more complex problem that also includes biology, behavioral science, etc. (33). Our findings may help identify individuals for whom grit assessment assists in obesity prevention (32).

The present study has several strengths. First, the inclusion of adult men and women across a wide range of age groups ensures the generalizability of our findings. Second, formal mediation analysis allowed us to quantify how multidimensional eating behaviors mediate the association between grit and obesity.

This study has several limitations. First, the possibility of reverse causality cannot be excluded due to the cross-sectional nature of the study. For example, a potential change in thoughts about eating behavior due to obesity (e.g., a strong preoccupation with the need for restraint due to obesity, etc.) would undermine the validity of our mediation analyses. Second, as in other studies (9), height and weight assessments were self-reported. Third, it is unclear whether the present findings on the interrelationships of obesity with grit and eating behaviors obtained in the Japanese food culture and a single ethnic society apply to other countries and racial groups.

In conclusion, a higher grit level was associated with a lower likelihood of obesity among Japanese adults across a wide range of age groups. Uncontrolled eating, emotional eating, and cognitive restraint mediated this association. Establishing a good and useful partnership between individuals and primary care physicians to identify separate impairments in eating behaviors, rather than focusing on an individual’s lack of willpower, may contribute to the prevention and management of obesity. However, these findings need to be validated by further research.

## Abbreviations

BMI: Body mass index
CR: Cognitive restraint
EE: Emotional eating
TFEQ-R21: Three-Factor Eating Questionnaire-R21
UE: Uncontrolled eating

## Acknowledgements

NK, TM, TA, and HK designed the research; NK, TM, and TW conducted the research; NK, JK, and HK analyzed the data; NK performed the statistical analyses; NK wrote the paper. NK was primarily responsible for the final content. All the authors have read and approved the final version of the manuscript.

## Data Availability*

The data described in this manuscript will be made available upon reasonable request from the corresponding author.

