## Supplementary Files for "The grit personality trait, eating behavior, and obesity among Japanese adults"

### Supplementary Item 1. Participant Sampling Strategy

Our panel survey recruited Japanese adults aged ≥20 years with the assistance of a web-based company (Cross Marketing, Shinjuku, Tokyo, Japan). The target sample size was 1500 participants due to the project's budget constraints. The sample size was set at a 1:1 ratio for men and women (750 each) and a 1:1 ratio for older and younger individuals of each sex (375 each for each sex). To sample individuals with and without obesity, sampling ratios were established using the original categories, for which the web-based company panel was registered in advance. Specifically, we set the number of individuals to ensure a 10:3:2 ratio among those who reported a history of hospitalization or clinic visits for obesity, those who were worried about obesity, and those who did not report these problems (i.e., the general population). (i.e., 250, 75, and 50 individuals for each sex and each age status, respectively).

**Supplementary Item 2. Designing Screener Items**

To assess the presence of careless participants, [[1]](https://paperpile.com/c/yiLwwt/rEPUf) five "screener" items were created to identify and exclude them from our analyses. Specifically, we excluded respondents with an inappropriate entry of 1) age or 2) gender or extreme values for 3) height or 4) weight, or those with 5) response time <5 min. [[2,3]](https://paperpile.com/c/yiLwwt/LGyDR+wMKeM)

Respondents were asked to indicate their age and sex twice, once at the beginning and once in the latter part of the questionnaire. We identified and excluded respondents whose two age or two sex responses did not match.

In the latter part of the questionnaire, the respondents were asked to indicate their height and weight.

Considering the realistic distribution of anthropometric characteristics among Japanese, we excluded those who reported their height to be either ≥210 cm or ≤120 cm. We also excluded those who reported their weight to be either ≥ 151 kg or ≤ 25 kg.

Based on our pilot test of the time required to complete the questionnaire, we excluded respondents who completed the survey in less than five minutes. Those who completed the questionnaire too quickly were categorized as careless [[1,2]](https://paperpile.com/c/yiLwwt/rEPUf+LGyDR) and were excluded from our analyses.

**Supplementary Table 1. Items and responses for The Three-Factor Eating Questionnaire Revised 21-Item (TFEQ-R21)**

Questionnaires in Japanese version are available from the following website (<https://noriaki-kurita.jp/wp-content/uploads/2024/04/84a60b0e2d5b33aa23a5f61c43ef8f46.pdf>)

| Instruction | Please circle the number for the response that best describes how much you agree or disagree with each of the following statements. |
| --- | --- |
| Question 1 | I deliberately take small helpings to control my weight. † |
| Question 2 | I start to eat when I feel anxious. † |
| Question 3 | Sometimes when I start eating, I just can’t seem to stop. † |
| Question 4 | When I feel sad, I often eat too much. † |
| Question 5 | I don’t eat some foods because they make me fat. † |
| Question 6 | Being with someone who is eating often makes me want to also eat. † |
| Question 7 | When I feel tense or ‘‘wound up’’, I often feel I need to eat. † |
| Question 8 | I often get so hungry that my stomach feels like a bottomless pit. † |
| Question 9 | My doctor is well qualified to manage (diagnose and treat or make an appropriate referral) medical problems like mine. † |
| Question 10 | When I feel lonely, I console myself by eating. † |
| Question 11 | I consciously hold back on how much I eat at meals to keep from gaining weight. † |
| Question 12 | When I smell a sizzling steak or see a juicy piece of meat, I find it very difficult to keep from eating - even if I've just finished a meal. † |
| Question 13 | I’m always hungry enough to eat at any time. † |
| Question 14 | If I feel nervous, I try to calm down by eating. † |
| Question 15 | When I see something that looks very delicious, I often get so hungry that I have to eat right away. † |
| Question 16 | When I feel depressed, I want to eat. † |
| Response options for Question 1 to 16 | Definitely true/Mostly true/Mostly false/Definitely false |
| Question 17 | How often do you avoid ‘‘stocking up’’ on tempting foods? |
| Response options for Question 17 | Almost never/Seldom/Usually/Almost always |
| Question 18 | How likely are you to make an effort to eat less than you want? |
| Response options for Question 18 | Unlikely/A little likely/Somewhat likely/Very likely |
| Question 19 | Do you go on eating binges even though you’re not hungry? |
| Response options for Question 19 | Never/Rarely/Sometimes/At least once a week |
| Question 20 | How often do you feel hungry? |
| Response options for Question 20 | Only at mealtimes/Sometimes between meals/Often between meals/Almost always |
| Question 21 | On a scale from 1 to 8, where 1 means no restraint in eating and 8 means total restraint, what number would you give yourself? Mark the number that best applies to you‡ |
| Response options for Question 21 | \| 1 \| 2 \| 3 \| 4 \| 5 \| 6 \| 7 \| 8 \| I eat whatever and whenever I want to I am constantly limiting my food intake, never "giving in" |

The original English version[[1]](https://paperpile.com/c/yiLwwt/WYRSU) is also provided for each item and response.

*The instructional statements generated through the formal process of translation are presented.

†Questions 1 through Question 16 are reverse-scored items.

‡Questions 21 was recoded as follows: scores 1–2 as 1, 3–4 as 2, 5–6 as 3, and 7–8 as 4.

Using the twenty-one questions, the following three domains can be assessed:

**Uncontrolled eating (UE)**: Question 3, Question 6, Question 8, Question 9, Question 12, Question 13, Question 15, Question 19, and Question 20.

**Cognitive restraint (CR)**: Question 1, Question 5, Question 11, Question 17, Question 18, and Question 21.

**Emotional eating (EE)**: Question 2, Question 4, Question 7, Question 10, Question 14, and Question 16

Before using this instrument, please register at https://noriaki-kurita.jp/resources/tfeq-r21-jpn/.

In addition, please cite this article as follows:

Kurita N, Maeshibu T, Aita T, Wakita T, Kikuchi H. The grit personality trait, eating behavior, and obesity among Japanese adults. *medRxiv.* 2024: 2024.04.13.24305766.

**Supplementary Item 3. Survey Items used for the analyses of the present study.**

The Japanese version of the Dutch Eating Behavior Questionnaire (DEBQ) is a 33-item instrument consisting of three factors: "emotional eating," which indicates eating behavior aroused by heightened emotions (13 items); "external eating," eating behavior triggered by external stimuli such as taste and smell (10 items); and, "restrained eating," the degree of intentional dietary restriction (10 items). [[1,2]](https://paperpile.com/c/yiLwwt/qxkoe+eUMPI) Respondents were instructed to rate each item on a Likert scale ranging from 1 (never) to 5 (very often). The average of the items was used as the score for each of the three domains. [[1]](https://paperpile.com/c/yiLwwt/qxkoe) Higher scores indicated that the construct was applied better. The alpha coefficients for each domain were as follows: emotional eating, 0.95; external eating, 0.73; and, restrained eating, 0.87. [[2]](https://paperpile.com/c/yiLwwt/eUMPI) Criterion-related validity of the Japanese version of the DBEQ was tested against body mass index in Japanese adults; although a weak negative correlation with external eating and a weak positive correlation with restrained eating were demonstrated, a correlation with emotional eating was not demonstrated. [[2]](https://paperpile.com/c/yiLwwt/eUMPI)

Demographic characteristics such as age, sex, education level, total household income, and marital status; healthy behaviors such as exercise habits, smoking history, and alcohol consumption; and, non-communicable diseases (diabetes, cancer, kidney disease, stroke, congestive heart failure, chronic lung disease, eating disorders, depression, and other mental disorders) were collected as covariates.

Sex was selected as either male or female. The educational level was junior high school, high school, professional training college, technology college, junior college, university, or graduate school. For the analyses, the data were collapsed into five levels (junior high school, high school, professional training college/college of technology/junior college, university, and graduate school). Total household income was classified as < 1,000,000 yen; 1,000,000 to < 3,000,000 yen; 3,000,000 to < 5,000,000 yen; 5,000,000 to < 10,000,000 yen; ≥ 10,000,000 yen. For the analyses, the data were collapsed into four levels (< 1,000,000 yen; 1,000,000 to < 5,000,000 yen; 5,000,000 to < 10,000,000 yen; ≥ 10,000,000 yen). Marital status was classified as unmarried, married, divorced, or widowed. The items on exercise habits, smoking, and alcohol consumption were adapted from a standard questionnaire for specific health checkups among Japanese adults as suggested by the Ministry of Health, Labor, and Welfare, Japan. [[3]](https://paperpile.com/c/yiLwwt/adzLH) Regular exercise was defined if the respondent chose "yes" from "yes" or "no" to the following question: "I do light, sweaty exercise of at least 30 minutes two days a week for at least one year. This question corresponds to ≥ 60 minutes per week of physical activity with an intensity ≥ 3 METS, which has been shown to reduce the risk of lifestyle-related diseases and mortality by 12%. [[3]](https://paperpile.com/c/yiLwwt/adzLH) Smoking was defined by a response of "yes" from "yes" or "no" to the following question: "Do you currently habitually smoke cigarettes?" The question was annotated so that "current habitual smokers" were defined as "those who smoked a total of ≥100 cigarettes or smoked for ≥6 months" and those who smoked during the last month. Alcohol consumption was chosen from one of the following three options in response to the question: " Frequency of alcohol consumption (sake, shochu, beer, western-style liquor, etc.)": every day, sometimes, rarely (cannot drink). For noncommunicable diseases, the following instructional statement was used: "Have you ever been told by a physician that you have any of the following diseases?" It was followed by the following nine items: diabetes mellitus, cancer, kidney disease, stroke, congestive heart failure, chronic lung disease, eating disorders, depression, and other mental disorders. For each item, one of three options was allowed: never told, told and visited a physician in the past, told and currently visiting a physician's office. If either of the latter two were chosen, the respondent was considered to have a non-communicable disease.

**Supplementary Item 4. Detailed description of statistical analyses**

Psychometric analyses were performed using R version 4.1.2, the psych package version 2.2.3, and the lavaan package version 0.6-11. All other analyses were performed using Stata/SE, version 17 (Stata Corp., College Station, TX, USA). Respondents' characteristics are summarized as means and standard deviations and 5th and 95th percentiles for continuous variables and as frequencies and proportions for categorical variables.

For the TFEQ-R21 and TFEQ-R18V2, three-factor confirmatory factor analyses were conducted to examine the goodness of fit of the three-factor model. Recoded raw scores were used (items 1–16 for the TFEQ-R21 and TFEQ-R18V2; item 21 for the TFEQ-R21). The goodness of fit of the models was assessed using the comparative fit index (CFI) and root mean square approximation error (RMSEA). Acceptable model criteria are a CFI ≥ 0.90 and an RMSEA ≤ 0.08.[[1]](https://paperpile.com/c/yiLwwt/YpRis) Acceptable standardized loadings were set at ≥ 0.3, while also considering the goodness of fit of the model and internal consistency reliability. [[2]](https://paperpile.com/c/yiLwwt/5Qc8A) The distributions of the responses were examined at the item level. Positive floor (>50% of the responses at the lower end of the possible scores) and ceiling effects (>50% of the responses at the upper end of the possible scores) were examined.  [[3]](https://paperpile.com/c/yiLwwt/WYRSU) The reliability of each domain in the three-factor models was assessed by Cronbach's α and McDonald's ω coefficients. [[4]](https://paperpile.com/c/yiLwwt/CYRR7) In general, coefficients > 0.7 are recommended. [[4]](https://paperpile.com/c/yiLwwt/CYRR7)

Furthermore, construct validity was examined by testing the correlations between the TFEQ domain scores and the Japanese version of the DEBQ and BMI. Spearman correlation coefficient was calculated to examine correlations.

**Supplementary Table 2. Three-factor structural validity and internal consistency reliability of the domains in the TFEQ-R21**

| **Goodness-of-fit indices based on confirmatory factor analysis** |  |
| --- | --- |
| Comparative Fit Index (CFI) | 0.924 |
| Root Mean Square Error of Approximation | 0.069 |
| Standardized Root Mean Square Residual | 0.059 |
| **Internal consistency reliability** |  |
| Cronbach's α coefficient |  |
| Uncontrolled Eating (UE) | 0.89 |
| Cognitive Restraint (CR) | 0.79 |
| Emotional Eating (EE) | 0.92 |
| McDonald's ω coefficient |  |
| Uncontrolled Eating (UE) | 0.89 |
| Cognitive Restraint (CR) | 0.78 |
| Emotional Eating (EE) | 0.93 |

### Supplementary Figure 1. Confirmatory factor analysis for TFEQ-R21

Dark gray squares indicate the items. Dark grey circles indicate domains. Digits overlaid on the one-way arrows indicate standardized loadings. The digits overlaid on the two-way arrows indicate the correlations.

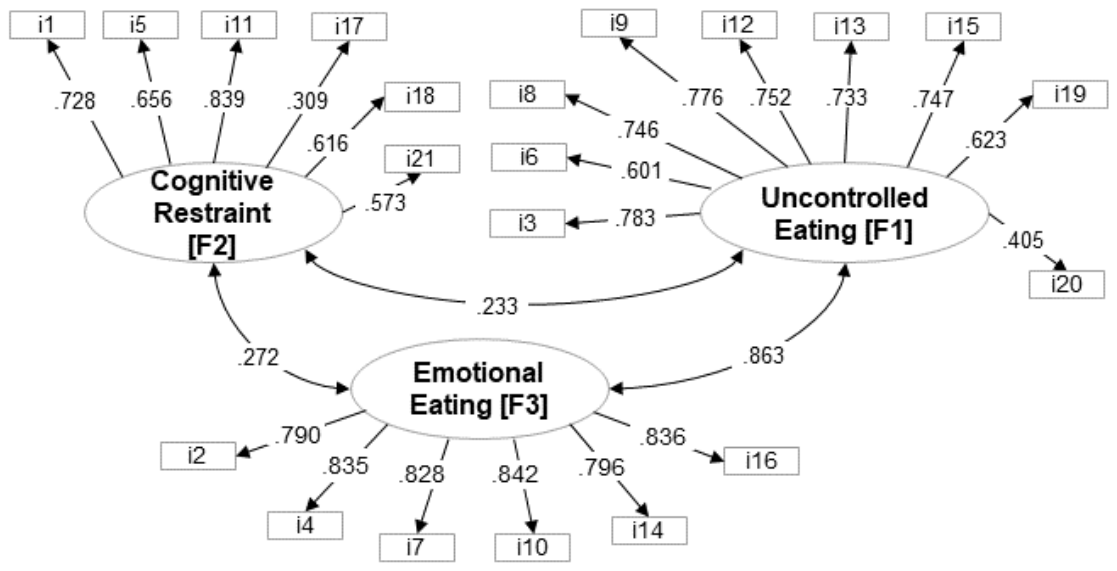

### Supplementary Table 3. Flooring and ceiling by item

| Total population (N = 1641) | | | | |  | Obesity subgroup (n = 439) | | | | |
| --- | --- | --- | --- | --- | --- | --- | --- | --- | --- | --- |
| % of flooring | |  | % of ceiling | |  | % of flooring | |  | % of ceiling | |
| Item | % |  | Item | % |  | Item | % |  | Item | % |
| 14R | 55.2 % |  | 6R | 7.3 % |  | 14R | 41.2 % |  | 6R | 12.1 % |
| 4R | 53.4 % |  | 17 | 7 % |  | 4R | 39.2 % |  | 3R | 10.5 % |
| 10R | 52.3 % |  | 11R | 5.9 % |  | 10R | 38.5 % |  | 8R | 7.7 % |
| 16R | 46.8 % |  | 21R | 5.5 % |  | 16R | 33.7 % |  | 17 | 7.3 % |
| 2R | 45.3 % |  | 18 | 5.3 % |  | 13R | 31.2 % |  | 7R | 7.1 % |
| 13R | 45.2 % |  | 3R | 5 % |  | 2R | 29.8 % |  | 2R | 6.6 % |
| 7R | 45 % |  | 5R | 4.9 % |  | 7R | 29.6 % |  | 19 | 5.9 % |
| 9R | 43.3 % |  | 8R | 4.3 % |  | 5R | 28 % |  | 5R | 5.7 % |
| 8R | 41.3 % |  | 7R | 4 % |  | 20 | 27.1 % |  | 11R | 5.5 % |
| 3R | 37.2 % |  | 2R | 3.8 % |  | 9R | 26.4 % |  | 4R | 5 % |
| 5R | 36.8 % |  | 1R | 3.3 % |  | 8R | 24.6 % |  | 15R | 4.8 % |
| 19 | 33.8 % |  | 4R | 3 % |  | 15R | 19.8 % |  | 16R | 4.3 % |
| 15R | 33.3 % |  | 19 | 3 % |  | 12R | 19.6 % |  | 18 | 4.3 % |
| 20 | 32.9 % |  | 15R | 2.8 % |  | 3R | 18.7 % |  | 10R | 4.1 % |
| 12R | 32.2 % |  | 16R | 2.6 % |  | 19 | 17.1 % |  | 12R | 4.1 % |
| 18 | 28.2 % |  | 12R | 2.6 % |  | 18 | 16.6 % |  | 9R | 3.6 % |
| 21R | 23.3 % |  | 10R | 2.4 % |  | 17 | 16.4 % |  | 20 | 3.6 % |
| 1R | 22.9 % |  | 20 | 2.3 % |  | 21R | 15.3 % |  | 21R | 3.6 % |
| 17 | 22.5 % |  | 9R | 2 % |  | 1R | 14.8 % |  | 1R | 2.7 % |
| 11R | 21.1 % |  | 14R | 1.7 % |  | 6R | 10 % |  | 14R | 2.5 % |
| 6R | 16.8 % |  | 13R | 1.7 % |  | 11R | 8.7 % |  | 13R | 2.3 % |

R indicates the re-coded score

Items 4, 10, and 14 are components of the emotional eating subdomain:

4. When I feel sad, I often eat too much.

10. When I feel lonely, I console myself by eating.

14. If I feel nervous, I try to calm down by eating.

**Supplementary Table 4. Criterion validity of the TFEQ-R21**

|  | DBEQ | | |  |
| --- | --- | --- | --- | --- |
|  | External eating | Restrained eating | Emotional eating | BMI |
| TFEQ-R21 |  |  |  |  |
| Uncontrolled eating (UE) | 0.6605; p <0.001 | 0.1155; p <0.001 | 0.6706; p <0.001 | 0.3173; p <0.001 |
| Cognitive restraint (CR) | 0.042; p =0.0889 | 0.7572; p <0.001 | 0.161; p <0.001 | 0.208; p <0.001 |
| Emotional eating (EE) | 0.505; p <0.001 | 0.1433; p <0.001 | 0.8029; p <0.001 | 0.2889; p <0.001 |

Spearman correlation coefficients between the three factors in the TFEQ-R21 and the three factors in the DBEQ and BMI are shown.

TFEQ-R21: The 21-item Three-Factor Eating Questionnaire, DEBQ: Dutch Eating Behaviour Questionnaire, BMI: body mass index.

### Supplementary Table 5. Decomposition of the association between grit and obesity into direct and indirect effects using the KHB-method† (N = 1641)

|  | **Eating behavior as one of multiple mediators** | | | | | | | |
| --- | --- | --- | --- | --- | --- | --- | --- | --- |
|  | **Uncontrolled eating** |  |  | **Cognitive restraint** |  |  | **Emotional eating** |  |
|  | Odds ratio, point estimate (95% CI) | P-value |  | Odds ratio, point estimate (95% CI) | P-value |  | Odds ratio, point estimate (95% CI) | P-value |
| Indirect effect |  |  |  |  |  |  |  |  |
| *Overall* | **0.75 (0.69 to 0.81)** | **<0.001** |  | 1.002 (0.95 to 1.05) | 0.938 |  | **0.84 (0.78 to 0.9)** | **<0.001** |
| *Eating behavior* | **0.78 (0.73 to 0.84)** |  |  | **1.05 (1.01 to 1.08)** |  |  | **0.87 (0.83 to 0.92)** |  |
| *Regular exercise* | **0.96 (0.92 to 0.99)** |  |  | **0.96 (0.93 to 0.99)** |  |  | **0.96 (0.93 to 0.992)** |  |
| *Smoking* | 1.001 (0.994 to 1.007) |  |  | 1 (0.998 to 1.003) |  |  | 1 (0.996 to 1.005) |  |
| *Alcohol* |  |  |  |  |  |  |  |  |
| *Rarely (cannot) drink* | Ref. |  |  | Ref. |  |  | Ref. |  |
| *Sometimes* | 0.99 (0.97 to 1.008) |  |  | 0.99 (0.97 to 1.008) |  |  | 0.99 (0.97 to 1.008) |  |
| *Everyday* | 1.01 (0.99 to 1.02) |  |  | 1.01 (0.993 to 1.02) |  |  | 1.01 (0.99 to 1.02) |  |
| Direct effect | 0.99 (0.79 to 1.24) | 0.937 |  | **0.73 (0.59 to 0.91)** | **0.005** |  | 0.88 (0.71 to 1.1) | 0.263 |
| Total effect | **0.74 (0.6 to 0.92)** | **0.007** |  | **0.73 (0.59 to 0.91)** | **0.004** |  | **0.74 (0.6 to 0.92)** | **0.006** |
| % of Total Effect mediated |  |  |  |  |  |  |  |  |
| *Overall* | 97% |  |  | -0.6% |  |  | 58.6% |  |
| *Eating behavior* | 80.7% |  |  | -14.4% |  |  | 44.5% |  |
| *Regular exercise* | 15.1% |  |  | 13.4% |  |  | 13.3% |  |
| *Smoking* | -0.2% |  |  | -0.1% |  |  | -0.1% |  |
| *Alcohol* |  |  |  |  |  |  |  |  |
| *Rarely (cannot) drink* | — |  |  | — |  |  | — |  |
| *Sometimes* | 3.1% |  |  | 3.1% |  |  | 3.1% |  |
| *Everyday* | -1.8% |  |  | -2.6% |  |  | -2.1% |  |

^†^The KHB method was used, which is derived from a linear latent variable model assumed to underlie the logit model and extend the decomposition properties of the linear model to the logit model. This allowed for the estimation of the overall, direct, and indirect effects in the logit model (obesity as the dependent variable; grit as the exposure variable; eating behavior, regular exercise, smoking, and alcohol as the mediator variables; and the other variables in Table 1 as covariates).
